# Global estimates of tuberculosis incidence during pregnancy and postpartum: a rapid review and modelling analysis

**DOI:** 10.1101/2025.06.20.25329850

**Authors:** Nyashadzaishe Mafirakureva, Anna Cartledge, Isobella Bradshaw, Adrie Bekker, Nicole Salazar-Austin, Sue-Ann Meehan, Landon Myer, Jasantha Odayar, Molebogeng X. Rangaka, Peter J. Dodd

## Abstract

**Introduction:** Despite known maternal, perinatal and infant health risks of tuberculosis during pregnancy, global estimates of incidence remain limited. Existing estimates are outdated, and do not include the postpartum period, HIV co-infection, age, or specific changes in risk, limiting our understanding of the true scale of disease in this understudied population.

**Methods:** We estimated the global incidence of tuberculosis among pregnant and postpartum women using a mathematical modelling approach. We leveraged age- and sex-stratified tuberculosis incidence data from the World Health Organization (WHO) and applied risk ratios derived from a systematic literature review of tuberculosis risk during pregnancy and the postpartum period.

**Findings:** Compared to non-pregnant women without HIV, tuberculosis incidence rate ratios (IRRs) were 1.34 (95% CI:1.1-1.54) during pregnancy and 1.91 (1.53-2.39) during postpartum among HIV-negative women. For women living with HIV, IRRs were 5.73 (2.64-10.94) during pregnancy and 3.58 (0.85-9.63) postpartum. We estimated 239,300 pregnant women (95% UI:216,100-262,100) and 97,600 postpartum women (90,000-105,200) developed tuberculosis disease globally in 2023, with HIV contributing to 21% and 11% of cases respectively. The WHO Southeast Asian Region had the highest incidence (115,900 pregnant [97,300-134,500]; 51,800 postpartum [45,700-57,800] cases), followed closely by the African Region (115,500 pregnant [101,600-129,400]; 42,700 postpartum [38,100-47,200] cases).

**Interpretation:** Pregnant and postpartum women face substantial tuberculosis risk yet remain underrepresented in global estimates. Our findings underscore the need for improved surveillance and targeted interventions to reduce tuberculosis incidence in this group.

**Funding:** MRC

## Introduction

Tuberculosis is a major public health threat, responsible for an estimated 1.25 million deaths in 2023.^1^ In many high burden settings, tuberculosis incidence peaks in women (used inclusively to refer to all people who may become pregnant, including those who do not identify as women) during the reproductive years (15-49 years).^2,3^ It has been suggested that pregnant and postpartum women are at increased risk of tuberculosis disease compared to non-pregnant women.^4^ Maternal tuberculosis is associated with adverse pregnancy outcomes, including low or very low birth weight, preterm birth, maternal mortality, stillbirth, and infant mortality.^5^ A high incidence of postpartum tuberculosis has been associated with increased maternal and infant mortality, particularly among people living with HIV (PLHIV).^6^ While tuberculosis preventive treatment (TPT) is not contraindicated in PLHIV, clinicians frequently exercise caution when prescribing chemoprophylaxis during pregnancy due to persistent safety concerns.^7^ Furthermore, diagnosing tuberculosis during pregnancy is challenging; tuberculosis symptoms (e.g. shortness of breath) may be similar to pregnancy symptoms, physiological changes in pregnancy may mask tuberculosis symptoms (e.g. gestational weight gain), and there may be hesitancy to screen pregnant women using chest radiographs.^7^

The true burden of tuberculosis among pregnant and postpartum women is not well understood. Tuberculosis during pregnancy is rarely reported separately, leaving substantial gaps in data and understanding. Pregnancy status is not typically recorded or routinely reported as a tuberculosis program indicator and tuberculosis data is not recorded in pregnancy registers.^8^ The only published modelling study to date, estimated that 216,500 pregnant women developed tuberculosis in 2011, with women in Africa accounting for 41% of this burden.^9^ No estimates were included for the postpartum period. Additionally, these estimates did not account for age-specific variations in tuberculosis incidence rates, or the elevated risk posed by pregnancy and postpartum periods, particularly in the context of HIV co-infection.

Since the publication of the only available global estimates in 2014, WHO estimates of tuberculosis incidence by age and sex have become available,^10^ and additional studies on the risk of tuberculosis during pregnancy have been published.^3^ This modelling study aims to update and expand estimates of tuberculosis incidence during pregnancy and the postpartum period relative to non-pregnant or non-postpartum periods. To do this, we first employed a rapid review and meta-analysis methodology to quantify the incidence rate ratios (IRRs) for tuberculosis during pregnancy and the postpartum period. Further, we sought additional data to quantify the risk of tuberculosis among pregnant and postpartum women living with HIV. Finally, we used age-specific tuberculosis incidence data and applied these IRRs within an established mathematical modelling framework to estimate global and country-specific tuberculosis incidence in pregnant and postpartum women by age and HIV infection status.

## Methods

A rapid review methodology was employed to identify and synthesise evidence on the risk of tuberculosis during pregnancy and postpartum. The research question was developed using a PECOS framework (Population, Exposure, Comparator, Outcome and Study). The review question was “Among women in the reproductive age group (Population), what is the relative risk of tuberculosis disease (Outcome) during pregnancy and the postpartum period (Exposure) compared to non-pregnant or non-postpartum women (Comparator), observed and reported in studies (Study)?” The protocol for the rapid review was registered with PROSPERO (identification number: CRD42018111690).

### Search Strategy

Two independent reviewers (AC and IB) conducted a systematic search of the Medline and EMBASE databases via OVID, without applying any date or language restrictions. The search aimed to capture studies at the intersection of terms related to “*pregnancy or postpartum*” as exposures, *“tuberculosis”* as the outcome, and “*observational study*” as the study design. The terms used for “*pregnancy or postpartum*” and *“tuberculosis”* were adapted from terms commonly applied in similar, relevant Cochrane reviews. For the term “*observational study*”, we applied filters recommended by the Scottish Intercollegiate Guidelines Network (SIGN)^11^ for observational studies.

To ensure comprehensive coverage, we supplemented the database search with hand-searching of the references and citations of the articles that were included after full-text review. An update of the Medline and EMBASE searches was conducted by a single reviewer (NM) in February 2024 to include the most recent evidence. We also utilised relevant professional networks to identify any data or studies that might have been missed during the search process.

### Study characteristics and inclusion criteria

The rapid review included three types of studies during full-text screening. Type 1 studies were longitudinal studies that reported tuberculosis incidence during pregnancy or postpartum, either with a non-pregnant control group or controlled internally through case series designs. Type 2 studies were cross-sectional studies reporting tuberculosis prevalence during pregnancy or postpartum, compared to a non-pregnant control group, and reporting odds ratios (ORs). Type 3 were cross-sectional studies reporting the prevalence of pregnancy or postpartum status in cohorts of individuals with tuberculosis, which reported ORs or provided data enabling the calculation of ORs. Type 3 studies were also included if post-hoc control group construction seemed feasible, based on publicly available population data on pregnancy rates. Studies were excluded if a suitable control group was not available or could not be reasonably constructed.

### Data Extraction and Analysis

Data extracted from studies selected after full-text screening included study location and period, study design, population characteristics, control group details, participant demographics (HIV status, age), tuberculosis screening and diagnostic approaches, tuberculosis case definition, proportion of cases with bacteriologic confirmation, and postpartum period definition. Discrepancies between reviewers (AC and IB) were addressed through discussion with a third independent reviewer (PJD) until consensus was reached. All data were cross-checked for accuracy and consistency. The NIH’s Quality Assessment Tool for Observational Cohort and Cross-Sectional Studies^12^ was used to assess the risk of bias in individual studies. The Preferred Reporting Items for Systematic Reviews and Meta-Analyses (PRISMA)^13^ guidelines were followed to report the rapid review findings. We provide a narrative summary and quality appraisal for the studies included. Studies rated as good were included in the quantitative synthesis.

### Meta-analysis

The primary outcome of the review was the incidence rate ratio (IRR) for tuberculosis during pregnancy and the postpartum period, compared to non-pregnant or non-postpartum women. Odds ratios (ORs) from cross-sectional studies were converted into IRRs to facilitate comparison, with a preference for adjusted ORs when available. Effect sizes from individual studies were pooled using inverse variance-weighting to generate summary IRRs. Heterogeneity between studies was assessed with I^2^ and τ^2^ statistics. Forest plots were generated to visualise the results.

### IRRs for PLHIV

A single analysis from the ongoing Obesogenic oRigins of maternal and Child metabolic Health Involving Dolutegravir (ORCHID) study (NCT 04991402),^14,15^ was available to quantify the risk of tuberculosis among pregnant and postpartum women living with HIV. We constructed aggregated person-time and incident tuberculosis event counts for women in the not-pregnant, pregnant and postpartum periods, and used these to compute rate ratios relative to not-pregnant.

### Modelling tuberculosis incidence during pregnancy and postpartum

The incidence of tuberculosis during pregnancy and postpartum was estimated using a combination of 2023 WHO data on age- and sex-disaggregated tuberculosis incidence,^16^ and United Nations World Population Prospects 2024^17^ estimates of population and fertility (Figure 1). The number of pregnancies was approximated using live birth estimates by maternal age (5-year age groups within 15–49 years) from the United Nations World Population Prospects 2024. To calculate person-time at risk, we estimated exposure separately for pregnancy and postpartum, using an average duration of 9 months for pregnancy and 3 months for postpartum.

**Figure 1.**
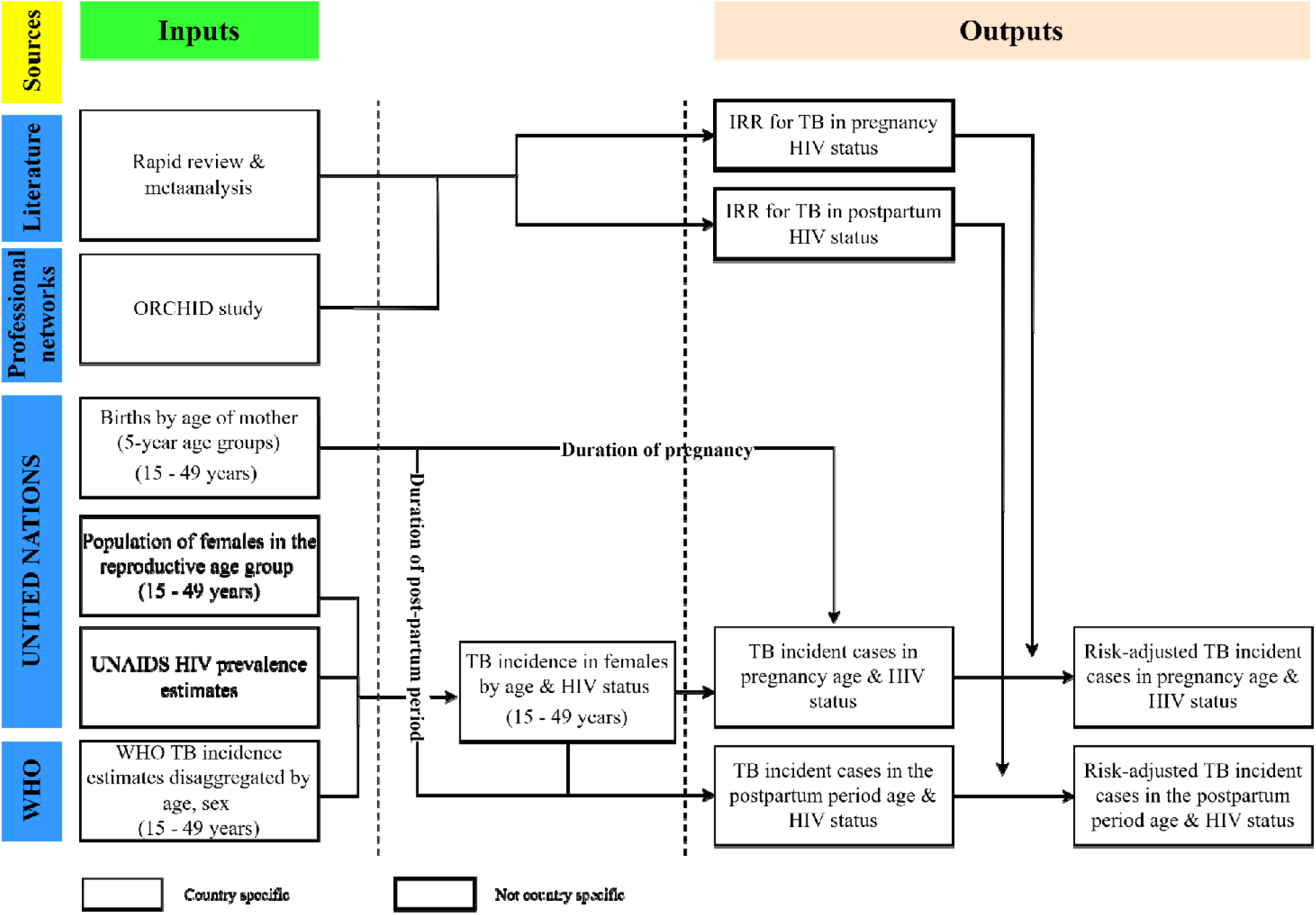
Methods model figure. HIV; human immunodeficiency virus, IRR; incidence rate ratio, ORCID; Obesogenic oRigins of maternal and Child metabolic Health Involving Dolutegravir study, TB; tuberculosis, WHO; World Health Organisation.

The baseline tuberculosis incidence rate in women of reproductive age was calculated using WHO tuberculosis incidence data and UN population estimates for females aged 15-49 years. This provided the annual tuberculosis incidence rate, which includes both women living with and without HIV. To account for the differential risk of tuberculosis by HIV status, the incidence rate was disaggregated for those living with HIV and those not living with HIV, using UNAIDS estimates of HIV prevalence among pregnant women and the relative risk of tuberculosis among women living with HIV. The relative risk of tuberculosis among women living with HIV (IRRs for tuberculosis given HIV) was estimated by comparing the odds of tuberculosis in HIV-positive women of reproductive age (derived from WHO tuberculosis burden data) and the odds of HIV in women of reproductive age (derived from UNAIDS HIV prevalence data), with adjustments for uncertainty and missing data. The disaggregated tuberculosis incidence estimates were then multiplied by the person-time at risk to estimate the total number of pregnant and postpartum women with active tuberculosis each year by HIV status.

Finally, to reflect the increased risk of tuberculosis associated with pregnancy and the postpartum period, we applied IRRs derived from our rapid review and meta-analysis. These IRRs, stratified by HIV status, adjusted the tuberculosis incidence estimates to capture the heightened vulnerability of women during pregnancy and postpartum.

Our analysis propagated uncertainty from both tuberculosis incidence estimates, HIV data, population and birth data, and IRRs (see Appendix for details). All results are reported as means with 95% uncertainty intervals (UIs).

## Results

### Study selection and assessment

The database search identified 2,035 records, of which 1,997 were excluded after deduplication and screening of titles and abstracts. Of 37 articles selected for a full-text review, 31 were excluded, leaving six articles for qualitative assessment (Appendix, Figure. 1). Most records (65%; 20/31) were excluded due to lack of a relevant control group. The six studies included in the qualitative assessment were published between 1996 and 2020, spanning an observation period from 1992 to 2014 (Table 1). These studies reported data from six countries: the Dominican Republic,^18^ Malawi,^19^ Mongolia,^20^ South Africa,^21^ Sweden,^4^ and the United Kingdom.^22^ The study designs comprised two case-control studies and four cohort studies. Two studies included data on HIV,^19,21^ with one focusing exclusively on HIV.^21^ Five studies reported data on both pregnancy and the postpartum period (defined as within six months after delivery),^4,18,19,21,22^ while one study focused exclusively on pregnancy.^20^ The quality of the six studies for our question was rated as poor (1 study),^19^ fair (1 study),^18^ and good (4 studies)^4,20–22^ based on the number of positive responses to the NIH’s Quality Assessment Tool questions.^12^ One additional study (ORCHID)^14,15^ from South Africa was found through professional networks.

**Table 1.** Summary of the six studies included in the qualitative review. AFB; Acid-Fast Bacilli, ANC; antenatal care, CXR; chest X-ray, HIV; human immunodeficiency virus, PCR: polymerase chain reaction, SD; standard deviation, TB; tuberculosis, UK; United Kingdom

| First author, year | Study years | Country | Population, setting | Study type | Control type | HIV status | Mean age and SD/range | TB case definition | Quality assessment |
| --- | --- | --- | --- | --- | --- | --- | --- | --- | --- |
| Crampin, 2004<sup>19</sup> | 1996-2001 | Malawi | TB patients of both genders identified via “enhanced passive surveillance” | Case control | Field based random sampling of the community | Cases: 68%+<br>Controls: 14%+ | No data given | Positive smear, culture or biopsy | Poor |
| Espinal, 1996<sup>18</sup> | 1992-1994 | Dominican Republic | Women aged 15-44 years | Case control | Recruited at the National Laboratory of Public Health when they presented for anonymous HIV testing | Cases: 5.7%+<br>Controls: 50%+ | 29 | Must have all of: typical signs and symptoms of TB, >1 smear of resp tract secretions + for AFB, no history of prior TB | Fair |
| Odayar, 2018<sup>21</sup> | 2013-2014 | South Africa | HIV-positive women during pregnancy and postpartum $\geq 18$ years at a large public sector primary health care facility that provides ANC to >4000 women annually | Retrospective cohort study | Women 18 months preconception | All + | 29 at enrolment<br>IQR 26-33 | Positive sputum smear microscopy, Xpert, TB culture, CXR abnormalities consistent with TB | Good |
| Rendell, 2016<sup>20</sup> | 2013 | Mongolia | Pregnant women diagnosed with TB from central TB dispensaries | Retrospective case study | General population | Population prevalence <0.1% | 27 (SD +/- 5.6, range 18-43) | Based on microbiological results or symptoms and CXR findings | Good |
| Zenner, 2012<sup>22</sup> | 1996-2008 | UK | All women with pregnancies that occur within the time period studies | Cohort study | Self-controlled case series | No data given | 29.5 (range 13-50) | Positive sputum culture, presence of clinical/radiological signs, symptoms | Good |
| Jonsson, 2022<sup>4</sup> | 2005-2013 | Sweden | Women of 15–49 years of age who had given birth in Sweden | Cohort study | Women neither pregnant nor postpartum | No data given | 15–49 years | Laboratory confirmation (culture or microscopy & PCR) and clinical/radiological findings | Good |

### IRRs

Four studies were included in the primary meta-analysis: Jonsson et al.^4^, Rendell et al.^20^, Zenner et al^22^ and Odayar et al. (2025)^15^ Odayar et al. (2018)^21^ was excluded as it focused exclusively on patients living with HIV and estimates were confounded by starting antiretroviral therapy and receiving tuberculosis preventive treatment. Crampin et al.^19^ and Espinal et al.^18^ were excluded from the primary synthesis because the quality was not rated as good for our purpose. The estimated summary IRR for tuberculosis among HIV negative women was 1.34 (95% CI: 1.1 - 1.54) during pregnancy and 1.91 (95% CI: 1.53 - 2.39) during the postpartum period compared to the non-pregnant and non-postpartum period (Figure 2). The IRR for tuberculosis among women living with HIV quantified from ORCHID,^14,15^ was 5.73 (95% CI: 2.64-10.94) during pregnancy and 3.58 (95% CI: 0.85-9.63) during the postpartum period (Figure 2).

**Figure 2.**
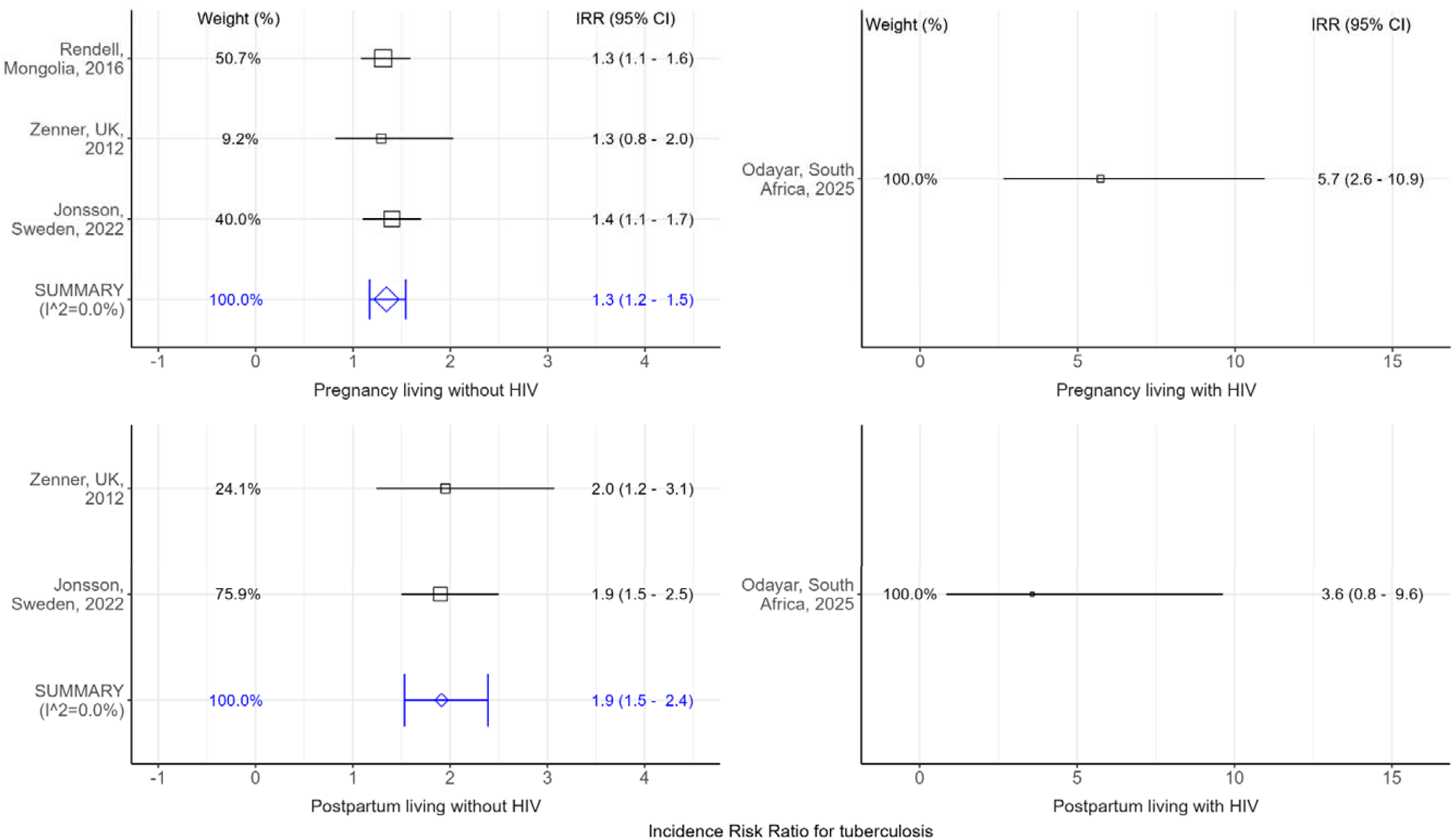
Incidence rate ratios (IRRs) for tuberculosis during pregnancy and postpartum, with and without HIV. CI; confidence interval, HIV; human immunodeficiency virus.

### Estimates of disease burden

The global estimated person-time at risk during 2023 was 101,158,100 person-years (95% UI: 97,160,600–105,155,700) during pregnancy and 32,966,700 person-years (95% UI: 31,663,900–34,269,500) during the postpartum period. An estimated 239,300 (95% UI: 216,100-262,100) pregnant women and 97,600 (95% UI: 90,000-105,200) women in the postpartum period developed active tuberculosis in 2023. The proportions of tuberculosis cases coinfected with HIV were 21% during pregnancy and 11% during postpartum. The highest burden was observed in the WHO Southeast Asian Region, with 115,900 (95% UI: 97,300, 134,500) pregnant women and 51,800 (95% UI: 45,700, 57,800) postpartum women affected (Table 2). The WHO African Region also carried a significant burden, with 115,500 (95% UI: 101,600, 129,400) pregnant women and 42,700 (95% UI: 38,100, 47,200) postpartum women having active tuberculosis. Across all WHO regional groupings, the majority of tuberculosis cases were observed in the 25-34 years age group (Figure 3).

**Table 2.** Global and regional estimates of tuberculosis incidence during pregnancy and postpartum disaggregated by HIV status. HIV; human immunodeficiency virus

|  | Mean (95% uncertainty interval) |  |  |  |  |  |
| --- | --- | --- | --- | --- | --- | --- |
| WHO region | Pregnancy |  |  | Postpartum |  |  |
|  | All pregnant women | Women living without HIV | Women living with HIV | All woman in the postpartum period | Women living without HIV | Women living with HIV |
| Africa | 115,500 (101,600, 129,400) | 73,600 (60,300, 87,000) | 41,900 (40,000, 43,700) | 42,700 (38,100, 47,200) | 34,100 (29,800, 38,500) | 8,500 (7,900, 9,100) |
| Americas | 5,100 (4,400, 5,700) | 3,800 (3,200, 4,500) | 1,200 (1,200, 1,300) | 2,000 (1,800, 2,200) | 1,800 (1,600, 2,000) | 200 (200, 300) |
| Europe | 1,100 (900, 1,400) | 900 (700, 1,200) | 200 (200, 200) | 500 (400, 500) | 400 (300, 500) | 0 (0, 100) |
| Southeast Asia | 115,900 (97,300, 134,500) | 108,300 (83,700, 132,900) | 7,600 (7,300, 7,900) | 51,800 (45,700, 57,800) | 50,200 (42,200, 58,200) | 1,500 (1,500, 1,600) |
| Western Pacific Region | 1,700 (1,300, 2,100) | 1,300 (800, 1,700) | 500 (400, 500) | 700 (500, 800) | 600 (400, 700) | 100 (100, 100) |
| Total | 239,300 (216,100, 262,500) | 187,900 (159,900, 215,900) | 51,400 (49,500, 53,300) | 97,600 (90,000, 105,200) | 87,200 (78,000, 96,300) | 10,500 (9,900, 11,100) |

**Figure 3.**
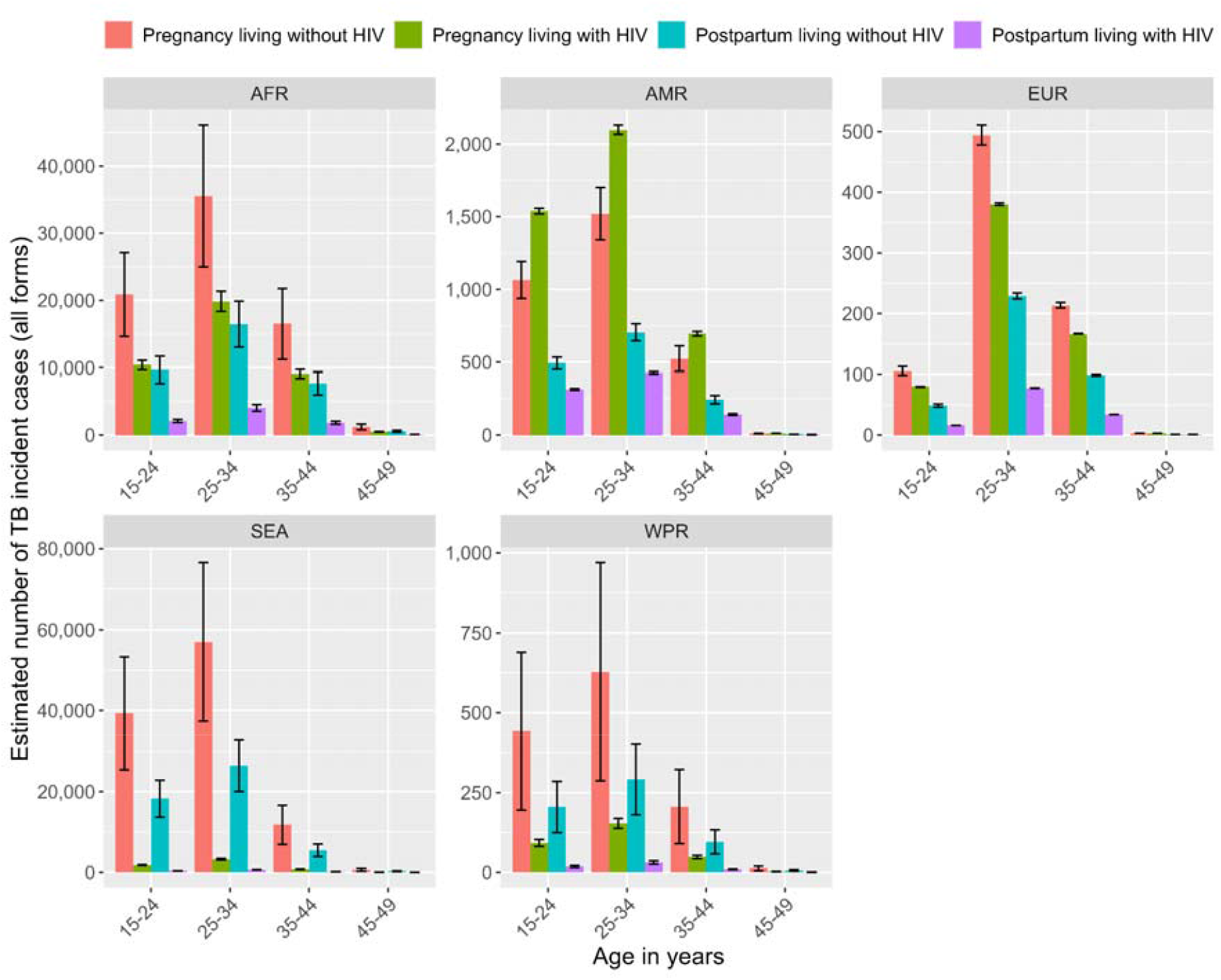
Age and HIV disaggregated tuberculosis incidence estimates during pregnancy and postpartum. AFR; Africa, AMR; Americas, EUR; Europe, SEA; Southeast Asia, WPR; Western Pacific Region, TB; tuberculosis.

## Discussion

This study responds to a critical evidence gap by providing the first updated global estimates of tuberculosis incidence during pregnancy and the postpartum period since the only previous analysis published a decade ago.^9^ Our findings further delineate the disproportionate burden attributable to HIV co-infection in this understudied population. Our literature review and meta-analysis provides evidence for increased risk of tuberculosis incidence during pregnancy and post-partum. Combining these IRRs with current estimates of age-specific fertility and tuberculosis incidence suggests a global tuberculosis incidence of 239,300 cases during pregnancy and 97,600 during the postpartum period (taken as 3 months), which in total represents 16% of tuberculosis incidence among women aged 15-44 years. These overall findings: i) add weight to the growing global consensus around the need to include pregnant and postpartum persons in research, including tuberculosis prevention trials, and ii) highlight the need to prioritise routine tuberculosis screening and testing among these women.

To put our global estimates in context, Cresswell et al.^23^ estimated a global mortality of 253□000 from maternal sepsis for the period 2009 - 2020. The incidence of tuberculosis in children is estimated to be more than 1 million globally,^1^ and the close contact between mothers and neonates is likely to be a substantial driver of tuberculosis incidence in the youngest children. We estimate 36% of tuberculosis incidence among pregnant and postpartum women in the WHO Africa region is among PLHIV, who are more likely to have particularly vulnerable children living with HIV.

Tuberculosis during pregnancy carries increased risks of premature birth, low birthweight, stillbirth and infant mortality, and vertical HIV transmission.^24^ Integrated, decentralized, and family-centred approaches to prevent, diagnose, and treat tuberculosis in pregnant women are part of the WHO Roadmap towards ending tuberculosis in children and adolescents, and pregnancy is specifically discussed in a number of WHO tuberculosis guidelines. Pregnancy is not considered a contraindication for women otherwise indicated for TPT, and the 6H regimen is recommended for all PLHIV, including pregnant women. In many scenarios, isoniazid-based TPT for PLHIV is expected to reduce combined maternal, fetal, and infant deaths, but more work is needed to predict risk and characterise alternate regimens.^25^ Diagnosis may be hampered by the WHO four-symptom screen having suboptimal sensitivity in pregnancy,^26^ and more work to optimize the integration of screening for tuberculosis disease into antenatal care is needed. Finally, data are lacking for this population on shorter treatment regimens, and in the case of drug-resistant tuberculosis.

Although previous studies have examined tuberculosis risk in pregnancy, our analysis provides some important advances. First, our work goes beyond a recent systematic review review,^3^ which independently identified the same evidence base but did not include a formal synthesis or HIV-specific risks, to produce quantitative estimates of risk during both pregnancy and the postpartum period. Second, our modelling advances upon the work by Sugarman et al. by: (1) encompassing the postpartum period as a distinct risk phase; (2) accounting for pregnancy- and post-partum associated changes in tuberculosis risk; (3) integrating HIV co-infection as an additional risk factor; and (4) employing age-disaggregated tuberculosis incidence data as a starting point. These methodological refinements likely explain why our 2022 estimate of 239□300 pregnancy-associated cases exceeds Sugarman’s 2011 projection (216□500 cases),^9^ despite an 8% decline in global tuberculosis incidence during this interval.^1^ This discrepancy suggests systematic underestimation in prior models that omitted HIV-confection and pregnancy-related risk elevation. Our approach enables stratification by maternal age and HIV status, which is important for planning prevention efforts in high-burden settings.

Our review identified only six studies with variable methodological quality and usefulness for estimating IRRs. While the two largest studies that used electronic health records for European populations showed concordant results,^4,22^ their generalisability to high-burden settings may be limited. Opportunities exist to expand this evidence base: the decade-old UK study^22^ could be replicated, and South Africa’s Western Cape Provincial Health Data Centre - a consolidation of all person level health data^27^ - could be used to generate comparable data in a high-incidence, HIV-endemic setting. Although one identified study in people living with HIV was unsuitable for IRR estimation due to potential confounding due to ART initiation,^21^ we derived HIV-specific IRRs from the ongoing ORCHID cohort.^14,15^

In using estimates of live births to estimate person time at risk, we did not account for multiple births (∼3% globally),^28^ stillbirths (∼1.4% of pregnancies),^29^ or abortions. While stillbirths and multiple births would likely have minimal impact on overall estimates, the effect of abortions remains unquantified due to scarce data on abortion timing and frequency, and a complete lack of evidence regarding whether tuberculosis incidence rate ratios apply equivalently to terminated pregnancies.

More importantly, we assumed that the same IRRs applied in all regions and regardless of age. The data on which our IRRs are based are dominated by cohorts from low-tuberculosis incidence European countries, and IRRs may differ elsewhere. IRRs of this magnitude are likely to capture both biological and social associations, and social associations between pregnancy and tuberculosis risk are more likely to vary regionally. More data from a variety of high tuberculosis-incidence settings would be useful to address this. While source studies used postpartum periods ranging from 3 to 6 months, we adopted a conservative 3-month duration for our definition of postpartum person-time. In effect, we assume that increases in tuberculosis risk seen in longer studies is concentrated in the first three months. Longer definitions of postpartum with these IRRs, or use of IRRs specific to the first 3 months postpartum, would each generate larger estimates of postpartum tuberculosis incidence.

Currently, modelling is required to estimate the incidence of tuberculosis in pregnant and postpartum women because direct data from current surveillance systems are lacking. Routinely capturing pregnancy status in tuberculosis registers and integrating tuberculosis screening into antenatal care in high-incidence settings would generate empirical data to validate and refine estimates while identifying patients in need of care - particularly in lower-income countries, where high fertility rates^30^ and tuberculosis prevalence during pregnancy intersect.^8^ These improvements, combined with registers that track pregnancy outcomes for TPT and anti-tuberculosis regimens, would strengthen the evidence base for safety profiles and programmatic decision-making. While modelled estimates remain critical for planning and advocacy, such real-world data would address pressing surveillance gaps and ensure policies reflect the realities of affected populations. Furthermore, systematically including pregnant women in research on TB screening, diagnosis, treatments, and vaccines is essential to generate evidence applicable to this group,^30,31^ closing the current inequity in intervention data and care delivery.

Pregnant and postpartum women are likely to be at increased risk of tuberculosis and large numbers in this group develop tuberculosis globally, affecting maternal health and neonatal and infant outcomes. More effort is needed to strengthen epidemiological and pharmacological surveillance, and to integrate tuberculosis screening in antenatal care.

## Data Availability

All code and data to reproduce this analysis are publicly available on GitHub https://github.com/petedodd/pregtb

https://population.un.org/wpp/

https://www.who.int/teams/global-programme-on-tuberculosis-and-lung-health/data

https://www.unaids.org/en

## Author contributions

PJD, AC and IB conceived and designed the study. AC, IB, NM and PJD conducted the systematic literature search and rapid review. LM and JA provided data to quantify the risk of tuberculosis among pregnant and postpartum women living with HIV. NM and PJD did the data analysis. NM, PJD and SAM wrote the first draft of the Article. All authors critiqued the results and revised and edited the submitted Article.

## Acknowledgements

PJD was supported by a fellowship from the UK Medical Research Council (MR/P022081/1); this UK-funded award is part of the EDCTP2 programme supported by the EU.

